# Perceptions and predictors of COVID-19 vaccine hesitancy among health care providers across five countries in sub-Saharan Africa

**DOI:** 10.1101/2022.10.11.22280952

**Authors:** Isabel Madzorera, Livesy Naafoe Abokyi, Edward Apraku, Temesgen Azemraw, Valentin Boudo, Christabel James, Dongqing Wang, Frank Mapendo, Ourohiré Millogo, Nega Assefa, Angela Chukwu, Firehiwot Workneh, Bruno Lankoande, Elena C. Hemler, Abbas Ismail, Sulemana Abubakari, Kwaku Poku Asante, Yemane Berhane, Japhet Killewo, Ayoade Oduola, Ali Sie, Abdramane Soura, Mary Mwanyika-Sando, Said Vuai, Emily Smith, Till Baernighausen, Raji Tajudeen, Wafaie W Fawzi

## Abstract

The African continent has some of the world’s lowest COVID-19 vaccination rates. While the limited availability of vaccines is a contributing factor, COVID-19 vaccine hesitancy among health care providers (HCP) is another factor that could adversely affect efforts to control infections on the continent. We sought to understand the extent of COVID-19 vaccine hesitancy among HCP, and its contributing factors in Africa. We evaluated COVID-19 vaccine hesitancy among 1,499 HCP enrolled in a repeated cross-sectional telephone survey in Burkina Faso, Ethiopia, Nigeria, Tanzania and Ghana. We defined COVID-19 vaccine hesitancy among HCP as self-reported responses of definitely not, maybe, unsure, or undecided on whether to get the COVID-19 vaccine, compared to definitely getting the vaccine. We used Poisson regression models to evaluate factors influencing vaccine hesitancy among HCP. Approximately 65.6% were nurses and the mean age (±SD) of participants was 35.8 (±9.7) years. At least 67% of the HCP reported being vaccinated. Reasons for low COVID-19 vaccine uptake included concern about vaccine effectiveness, side effects and fear of receiving unsafe and experimental vaccines. COVID-19 vaccine hesitancy affected 45.7% of the HCP in Burkina Faso, 25.7% in Tanzania, 9.8% in Ethiopia, 9% in Ghana and 8.1% in Nigeria. Respondents reporting that COVID-19 vaccines are very effective (RR:0.21, 95% CI:0.08, 0.55), and older HCP (45 or older vs.20-29 years, RR:0.65, 95% CI: 0.44,0.95) were less likely to be vaccine-hesitant. Nurses were more likely to be vaccine-hesitant (RR 1.38, 95% CI: 1.00,1.89) compared to doctors. We found higher vaccine hesitancy among HCP in Burkina Faso and Tanzania. Information asymmetry among HCP, beliefs about vaccine effectiveness and the endorsement of vaccines by the public health institutions may be important. Efforts to address hesitancy should address information and knowledge gaps among different cadres of HCP and should be coupled with efforts to increase vaccine supply.

## INTRODUCTION

The Coronavirus disease 2019 (COVID-19) caused by the novel SARS-CoV-2 continues to be a major public health challenge on the African continent and globally. As of March 8, 2022, there were at least 433 million cases of COVID-19 worldwide, and over 5.9 million associated deaths (1). On the African continent, close to 8.4 million cases and 170,300 deaths had been recorded, accounting for 3% of all global COVID-19 related mortality in the same period (1). COVID-19 however could continue to pose a threat to communities in sub-Saharan Africa (SSA) in the future, largely due to the lower availability of vaccines and hesitancy to accept vaccines by some in the population (2).

While other regions have made significant investments in vaccine rollout to curtail the further spread of COVID-19, efforts on the African continent continue to be affected by global inequity in access to vaccines (3). By the end of December 2021, only 7 African countries had achieved the target of vaccinating 40% of their population (4). While at least 68% of the global population has received at least one dose of the COVID-19 vaccine, Africa has the lowest share of the global vaccinated population with approximately 22% of the population having been fully vaccinated, and less than 1% have received a booster dose against COVID-19 (5, 6). Among African countries, coverage of at least one dose of vaccine ranged from 86% in Seychelles, 80% in Mauritius, 80% in Rwanda, 70% in Comoros and 69% in Botswana, to less than 6% in Cameroon, Madagascar, Burundi and Democratic Republic of Congo (5, 6). The potential undeterred spread of the COVID-19 pandemic and potential future mutations has serious ramifications for a continent already dealing with significant health and economic challenges, including food insecurity, high food prices, inadequate diets, and slowed economic growth (7, 8).

COVID-19 vaccine hesitancy is a global threat to achieving herd immunity (9). On the African continent, only 1 in 4 health workers is fully vaccinated against COVID-19 (10). The low levels of vaccination among health workers in Africa may be due to the lower availability of COVID-19 vaccines but also partly influenced by a hesitancy to take the vaccine. COVID-19 vaccine hesitancy among health care providers could be a serious threat to efforts to combat the pandemic in Africa as health workers play an essential role in the management and control of COVID-19 and have a high risk of getting infected. They are also a source of information on COVID-19 for the public and exert influence on public opinion in their contexts. COVID-19 vaccine hesitancy among healthcare workers could contribute to hesitancy in the general public and increase patient risk of contracting COVID-19.

Vaccine hesitancy, defined as the delay or reluctance of people or communities to receive safe and recommended vaccines, predates the COVID-19 pandemic (11, 12). Early studies in some SSA countries indicated a high willingness to take COVID-19 vaccines, however, this was before the availability of the vaccines (11, 13). A few studies have assessed hesitancy in COVID-19 vaccine uptake among health workers in Africa (9, 14, 15). A systematic review found COVID-19 acceptance rates of 46% across Africa and elevated levels of vaccine hesitancy have been reported (16, 17). However, acceptance rates as low as 28% have been reported in Central Africa (16). Factors influencing attitudes towards COVID-19 vaccines include fears about vaccine safety given the rapid development of vaccines, serious side effects, efficacy, lack of information and distrust of science and religious reasons (16). Gaps remain in our understanding of the extent of vaccine hesitancy among HCP in various contexts in SSA and the factors associated with it. This study contributes to understanding these gaps.

Understanding vaccine hesitancy and its predictors among HCP is important to inform strategies to enhance vaccination rates on the African continent (11). This study aimed to assess the magnitude and determinants of COVID-19 vaccine hesitancy among HCP across five countries in SSA that are part of the Africa Research Implementation Science and Education (ARISE) Network, Burkina Faso, Ethiopia, Nigeria, Tanzania and Ghana.

## MATERIAL AND METHODS

### Study setting

The study was part of a repeated cross-sectional survey conducted to assess knowledge and practices related to COVID-19 prevention and vaccination, and to evaluate the impact of COVID-19 on nutrition, health and other domains among adolescents, adults and HCP in 5 countries included in the ARISE Network. The ARISE network is a platform for public health research and training and includes 21 member institutions across nine sub-Saharan African countries.

Study participants for this study were HCP currently employed in health centres in the study areas. Briefly, in the baseline survey, we collected data from HCPs in urban areas of 3 SSA countries, Burkina Faso (Ouagadougou), Ethiopia (Addis Ababa), and Nigeria (Lagos). In round 2 of the survey, data were collected from these sites and additionally in rural Ghana (Kintampo) and urban Tanzania (Dar es Salaam). Further information on the ARISE sites in Burkina Faso, Ethiopia and Nigeria and their characteristics are described elsewhere (18, 19). Information on the design of the round 2 survey is described on Harvard University Center for African Studies website (https://africa.harvard.edu/files/african-studies/files/arise_covid_survey_round_2_methods_brief_final.pdf).

In the new round 2 sites of Tanzania and Kintampo, the study obtained HCP lists from medical professional associations and healthcare facilities in urban Dares Salaam and rural Kintampo. In all sites, the inclusion criteria were HCPs currently working in a healthcare setting, inclusive of all types of health facilities where COVID-19-related services were provided. We excluded dentists, pharmacists and other health providers in specialties unlikely to deliver COVID-19 related medical services. Fig 1 shows the sites included in the study.

**Figure 1:**
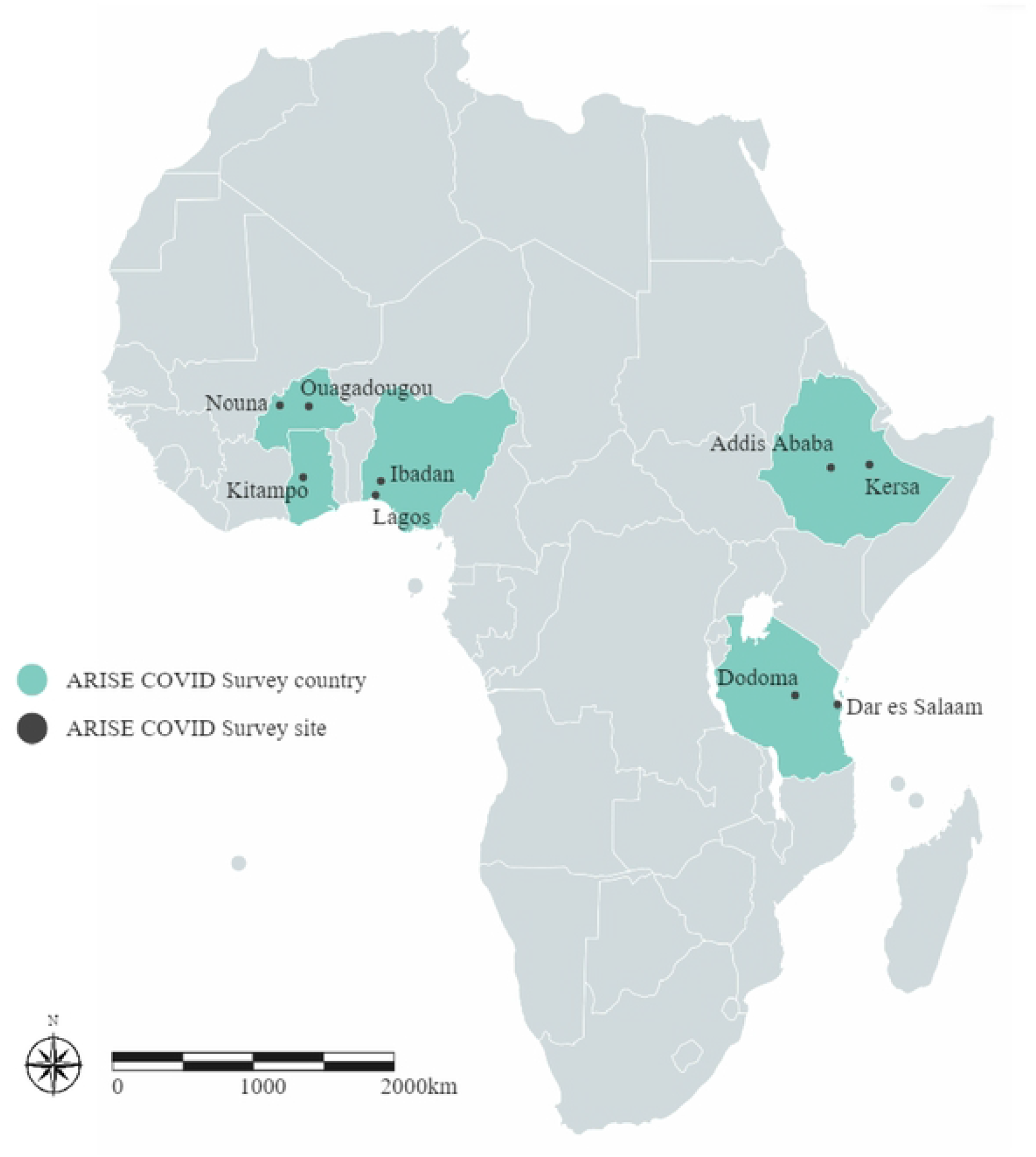
Map of study sites included in the ARISE COVID-19 Round 2 Survey, 2021.

### Study design

This study utilized computer-assisted telephone interviews to collect data from HCP currently employed in government, public and private health facilities in the study sites. Sampling frames for the study were developed using databases of HCP and their telephone numbers provided by professional associations and health facilities in each country. The study sites randomly selected 500 HCP to interview from the provided sampling lists in each country in Round 1, with a target to recruit 300 HCP. The target sample size for the round 2 survey was 300 HCP from each of the sites in the study. In round 2, participants from round 1; in Ethiopia, Burkina Faso and Nigeria, who were available were first interviewed. New participants were then randomly recruited from existing sampling frames to replace unavailable participants in round 1 to meet study sample size requirements.

Study data were collected by trained research assistants using standardized survey questionnaires that were adopted to the sites. Research assistants collected data on socio-demographic characteristics, including age, sex, occupation of the HCP, their knowledge, attitudes, practices and perceptions of COVID-19, as well as vaccine-related beliefs and hesitancy. Data collection was conducted between July to December of 2021. Round 1 survey data collection was conducted from July to November of 2020. Of the 900 HCPs interviewed in round 1, there were 548 participants retained in round 2 of the survey indicating a retention rate of 61%.

### Outcome: Vaccine hesitancy

We asked respondents if a COVID-19 vaccine were available, would they get it. We defined COVID-19 vaccine hesitancy among unvaccinated HCPs as responses of definitely not getting the vaccine, maybe, unsure, or undecided on whether to get the COVID-19 vaccine were it available, compared to responses of definitely getting the vaccine. If the HCP was already vaccinated, they were classified as not vaccine-hesitant. Based on these survey responses, we created a binary variable for COVID-19 vaccine hesitancy (Yes/No).

### Statistical analysis

We used descriptive and inferential statistics for the analysis. Descriptive statistics used frequencies for categorical variables and means and standard deviations for continuous variables to summarize socio-demographic characteristics, perceptions around COVID-19 vaccines, workplace practices and key COVID-19 related practices in round 2 of the survey. We used generalized estimating equation (GEE) Poisson regression models to evaluate associations between sociodemographic and other characteristics with vaccine hesitancy among HCPs in Round 2 of the study.

We evaluated factors associated with vaccine hesitancy among HCP. We considered the following possible predictors of vaccine hesitancy: age (20-29, 30-39, ≥ 40 years); respondent sex (female/male); occupation (doctor, nurse, other), health facility (government facility, private hospital, heath outpost or other), religion (Catholic or none Muslim, orthodox Christian, Protestant or other), self-perceived risk of COVID-19 exposure (no risk, low risk, very high risk, high risk), perceived effectiveness of COVID-19 vaccine (not effective at all, not very effective, somewhat effective, very effective), perceived safety of COVID-19 vaccines (very safe, somewhat safe, neither safe nor unsafe, not very safe, not at all safe), COVID-19 testing available in the facility where the HCP worked (Yes/No, free testing/paid testing). We also considered the type of COVID-19 testing available (Antigen test, PCR), having tested positive for COVID-19 previously (Yes/No), having cared for COVID-19 patients previously (never, yes and in the past one month, yes but over one month ago), workplace COVID-19 polices (Yes/No), influence of vaccine production in Africa on willingness to take vaccine (No, will not change my mind/Yes, will decrease my chances of taking it/Yes, will increase my chances of taking it), COVID 19 control practices (score indicating level of prevention measures being implemented in the workplace including wearing masks, using personal protective equipment (PPE), hand washing with water and soap, social distancing, sanitizers or hand washing station in health facility, cleaning and decontamination or disinfection of public areas, checking high temperatures, Yes/No), believe COVID-19 vaccine is bioweapon (Yes/No) and World Health Organization (WHO)/UNICEF endorsement of COVID-19 vaccine as safe and effective affect likelihood get the vaccine (much more likely, more likely, no difference, less likely and much less likely)

Further, we also considered a score of reasons for not getting the COVID-19 vaccine (Do not think it is needed, not at risk of getting COVID, vaccine not effective against COVID-19, negative media reports, vaccine not safe/ developed too fast, concerned about side effects, fear of experimental vaccine, will get worse quality vaccines, fear getting COVID-19 disease from the vaccine, illnesses/autism from the vaccine, will cause infertility/sterilization/population control, religious reasons/church, microchipping fears, New World Order, bad reaction with previous vaccinations, chronic condition e.g. diabetes, hypertension and personal liberty/do not want bodily intrusion.

Covariates were selected for inclusion in the main model using univariate tests at p<0.20. We evaluated for significant associations in the adjusted models based on a significance level of p<0.05. We used the missing indicator approach to account for missing covariate data. Analysis was conducted using SAS 9.4 (Cary, NC, USA).

### Ethical approval and consent

Verbal consent was obtained from study participants before they were admitted into the study. Ethical approval for the study was obtained from the Institutional Review Board at Harvard T.H. Chan School of Public Health and ethical review boards in each country and site, including the Kintampo Health Research Centre Institutional Ethics Committee in Ghana; the Nouna Health Research Center Ethical Committee and National Ethics Committee in Burkina Faso; the Institutional Ethical Review Board of Addis Continental Institute of Public Health in Ethiopia; the University of Ibadan Research Ethics Committee and National Health Research Ethics Committee in Nigeria; and the Muhimbili University of Health and Allied Sciences and National Institute for Medical Research in Tanzania.

## RESULTS

There were 1499 HCP included in the study across the 5 SSA countries. Of the participants, 300 were from Burkina Faso, 277 were from Ethiopia, 312 were from Nigeria, 310 were from Tanzania and 300 were from Ghana. Table 1 shows the socio-demographic characteristics of the study population. The majority of respondents (59.8%) across all countries were female. In Burkina Faso, however, female participants were fewer (46%) compared to men. The mean age (±SD) of respondents was 35.8 (±9.7) years. Most HCP in Ghana and Ethiopia were under 30 years of age, while in all other countries most respondents were in the 30-44 years age group. The majority of HCP assessed across all the countries were nurses (65.6%), with Ghana and Burkina Faso accounting for the highest proportion of nurses with 89.7% and 65.7% respectively. The majority of respondents worked in government hospitals or clinics. In Tanzania, 41.3% of HCPs worked in health posts where primary health care services were provided, while 24.3% from Ghana worked in Mission hospitals. In Ethiopia and Nigeria, most of the respondents reported Orthodox Christian religion. In Burkina Faso 49.8% were Catholic, and in Ghana and Tanzania at least 48% were Protestant or from other Christian denominations.

**Table 1:**
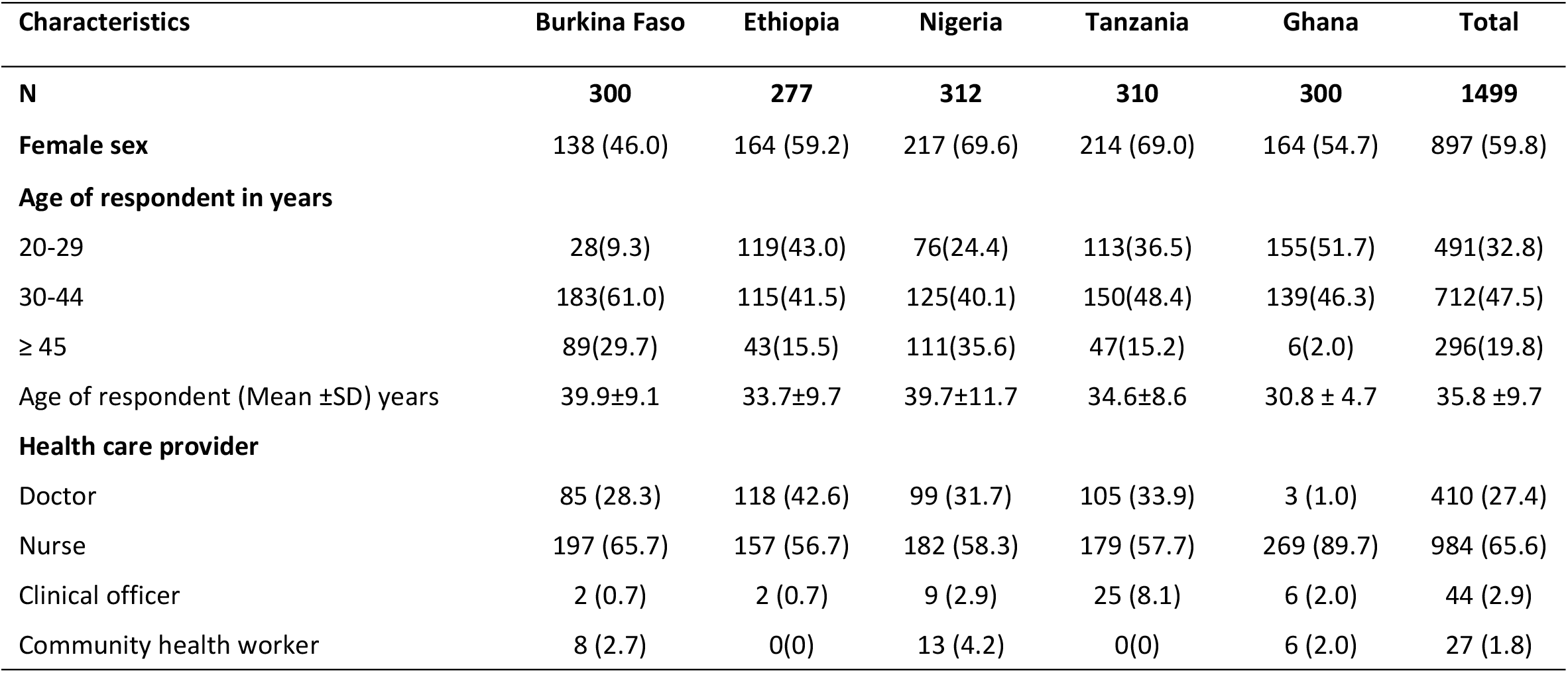
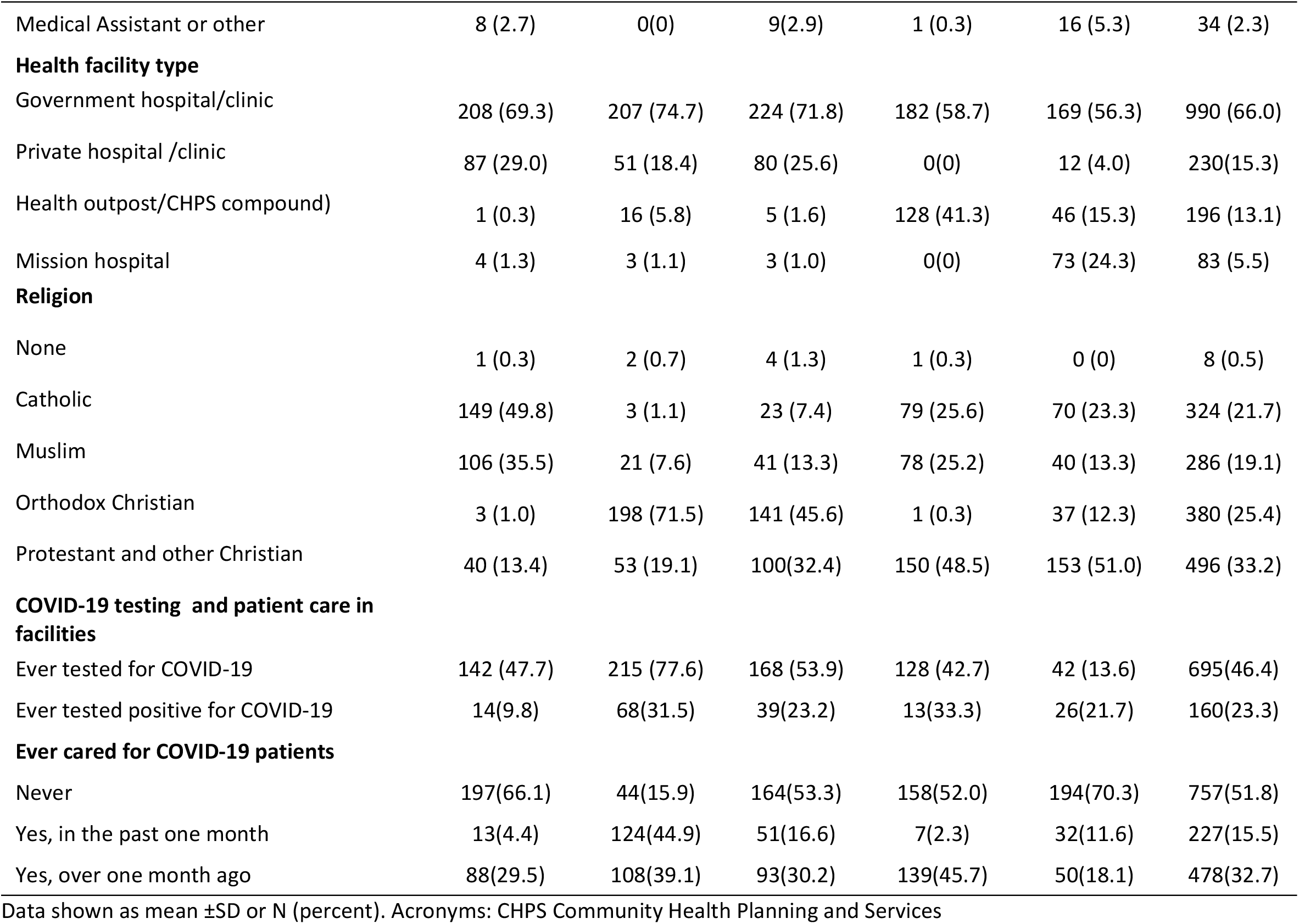
Socio-demographic characteristics of HCP across 5 countries in sub-Saharan Africa.

| Characteristics | Burkina Faso | Ethiopia | Nigeria | Tanzania | Ghana | Total |
| --- | --- | --- | --- | --- | --- | --- |
| **N** | **300** | **277** | **312** | **310** | **300** | **1499** |
| **Female sex** | 138 (46.0) | 164 (59.2) | 217 (69.6) | 214 (69.0) | 164 (54.7) | 897 (59.8) |
| **Age of respondent in years** |  |  |  |  |  |  |
| 20-29 | 28(9.3) | 119(43.0) | 76(24.4) | 113(36.5) | 155(51.7) | 491(32.8) |
| 30-44 | 183(61.0) | 115(41.5) | 125(40.1) | 150(48.4) | 139(46.3) | 712(47.5) |
| $\geq 45$ | 89(29.7) | 43(15.5) | 111(35.6) | 47(15.2) | 6(2.0) | 296(19.8) |
| Age of respondent (Mean ±SD) years | 39.9±9.1 | 33.7±9.7 | 39.7±11.7 | 34.6±8.6 | 30.8 ± 4.7 | 35.8 ±9.7 |
| **Health care provider** |  |  |  |  |  |  |
| Doctor | 85 (28.3) | 118 (42.6) | 99 (31.7) | 105 (33.9) | 3 (1.0) | 410 (27.4) |
| Nurse | 197 (65.7) | 157 (56.7) | 182 (58.3) | 179 (57.7) | 269 (89.7) | 984 (65.6) |
| Clinical officer | 2 (0.7) | 2 (0.7) | 9 (2.9) | 25 (8.1) | 6 (2.0) | 44 (2.9) |
| Community health worker | 8 (2.7) | 0(0) | 13 (4.2) | 0(0) | 6 (2.0) | 27 (1.8) |
| Medical Assistant or other | 8 (2.7) | 0(0) | 9(2.9) | 1 (0.3) | 16 (5.3) | 34 (2.3) |
| **Health facility type** |  |  |  |  |  |  |
| Government hospital/clinic | 208 (69.3) | 207 (74.7) | 224 (71.8) | 182 (58.7) | 169 (56.3) | 990 (66.0) |
| Private hospital /clinic | 87 (29.0) | 51 (18.4) | 80 (25.6) | 0(0) | 12 (4.0) | 230(15.3) |
| Health outpost/CHPS compound) | 1 (0.3) | 16 (5.8) | 5 (1.6) | 128 (41.3) | 46 (15.3) | 196 (13.1) |
| Mission hospital | 4 (1.3) | 3 (1.1) | 3 (1.0) | 0(0) | 73 (24.3) | 83 (5.5) |
| **Religion** |  |  |  |  |  |  |
| None | 1 (0.3) | 2 (0.7) | 4 (1.3) | 1 (0.3) | 0 (0) | 8 (0.5) |
| Catholic | 149 (49.8) | 3 (1.1) | 23 (7.4) | 79 (25.6) | 70 (23.3) | 324 (21.7) |
| Muslim | 106 (35.5) | 21 (7.6) | 41 (13.3) | 78 (25.2) | 40 (13.3) | 286 (19.1) |
| Orthodox Christian | 3 (1.0) | 198 (71.5) | 141 (45.6) | 1 (0.3) | 37 (12.3) | 380 (25.4) |
| Protestant and other Christian | 40 (13.4) | 53 (19.1) | 100(32.4) | 150 (48.5) | 153 (51.0) | 496 (33.2) |
| **COVID-19 testing and patient care in facilities** |  |  |  |  |  |  |
| Ever tested for COVID-19 | 142 (47.7) | 215 (77.6) | 168 (53.9) | 128 (42.7) | 42 (13.6) | 695(46.4) |
| Ever tested positive for COVID-19 | 14(9.8) | 68(31.5) | 39(23.2) | 13(33.3) | 26(21.7) | 160(23.3) |
| **Ever cared for COVID-19 patients** |  |  |  |  |  |  |
| Never | 197(66.1) | 44(15.9) | 164(53.3) | 158(52.0) | 194(70.3) | 757(51.8) |
| Yes, in the past one month | 13(4.4) | 124(44.9) | 51(16.6) | 7(2.3) | 32(11.6) | 227(15.5) |
| Yes, over one month ago | 88(29.5) | 108(39.1) | 93(30.2) | 139(45.7) | 50(18.1) | 478(32.7) |

Data shown as mean ± SD or N (percent). Acronyms: CHPS Community Health Planning and Services

Testing services were available for 85% of the respondents in Ethiopia, 67% in Nigeria, 59% in Burkina Faso, 40% in Ghana and 38% in Tanzania (results not shown). Among those facilities that had COVID testing services, most of the services were free. COVID-19 testing was free for the majority of respondents in Burkina Faso (98%), Ethiopia (90%), Nigeria (81%) and Ghana (98%). COVID-19 testing was paid for 60% in Tanzania and 19% in Nigeria, and less than 10% of the time in the other countries. Availability of PCR and antigen testing for COVID-19 was comparable, with respondents reporting 28.6% availability for the former compared to 25.4% for the latter. Of those HCP who had ever tested for COVID-19, about 23.3% had tested positive (Table 1). Approximately 52% of HCP in the study had never cared for a COVID-19 patient, and 15.5% had cared for patients affected in the previous month before the survey.

Table 2 shows HCP prevention measures implemented in the workplace and beliefs around COVID-19. The measures that most HCPs reported as being implemented in the workplace were wearing a mask (97.4%), handwashing with water and soap (97.6%), and regular cleaning or decontamination of public areas (92.0%). The least commonly practiced measures were wearing PPE (73.3%) and socially distancing patients in waiting rooms (69.9%). The use of personal protective equipment (PPE) was least reported in Ghana (55.2%) and Tanzania (62.3%). In Burkina Faso, socially distancing patients was reported by only 44.4% of HCP, and by 65.7% in Tanzania. In Ethiopia, temperature checks were reported by only 55.6% and in Tanzania by 67.1%.

**Table 2:** Frequency of COVID-19 preventive practices and beliefs about vaccines among HCP across 5 countries in sub-Saharan Africa.

| COVID-19 preventive practices and beliefs about vaccines | Burkina Faso | Ethiopia | Nigeria | Tanzania | Ghana | Total |
| --- | --- | --- | --- | --- | --- | --- |
| **COVID-19 Prevention practices** |  |  |  |  |  |  |
| Wearing masks | 295(98.3) | 275(99.3) | 311(100) | 279(90.3) | 298(99.3) | 1458(97.4) |
| Using personal protective equipment (PPE) such as gowns, goggles, shields, etc. | 274(91.6) | 231(83.7) | 231(74.8) | 192(62.3) | 165(55.2) | 1093(73.3) |
| Hand washing with water and soap | 296(98.7) | 258(93.1) | 309(99.4) | 299(96.5) | 300(100) | 1462(97.6) |
| Keeping sufficient distance between patients in the waiting area | 131(44.4) | 183(66.3) | 280(90.3) | 203(65.7) | 243(81.5) | 1040(69.9) |
| Presence of sanitizers or hand washing station in all service delivery points | 268(89.9) | 208(75.6) | 301(97.1) | 303(97.7) | 293(98.0) | 1373(92.0) |
| Regular cleaning and decontamination/disinfection of public areas and offices | 268(89.3) | 220(80.3) | 286(93.2) | 241(77.7) | 292(97.3) | 1307(87.7) |
| Temperature measurement (high temperatures 37 degrees Celsius) | 257(86.0) | 154(55.6) | 279(89.7) | 208(67.1) | 288(96.0) | 1186(79.2) |
| **Beliefs about COVID-19 vaccines** |  |  |  |  |  |  |
| Vaccine trial participants have died after taking the vaccine | 37(19.6) | 51(22.4) | 53(23.7) | 20(9.5) | 26(10.6) | 187(17.1) |
| Side effects from the COVID-19 vaccine are usually mild and temporary, and should go away in a few days | 253(94.4) | 223(80.8) | 261(86.4) | 216(83.4) | 278(95.5) | 1231(88.2) |
| There is no need for a vaccine because COVID-19 is a conspiracy or a bioweapon | 26(10.4) | 16(6.1) | 10(3.4) | 8(2.9) | 12(4.3) | 72(5.3) |
| People on the African continent are immune to COVID-19, so there is no need for a vaccine | 13(4.9) | 17(6.2) | 5(1.7) | 10(3.4) | 12(4.1) | 57(4.0) |
| It is not necessary to get a COVID-19 vaccine if you follow all safety protocols | 56(19.7) | 32(11.6) | 16(5.3) | 10(3.3) | 23(7.8) | 137(9.4) |
| There is not enough evidence that the COVID-19 vaccine prevents the occurrence and spread of COVID-19 | 166(65.6) | 89(32.7) | 143(49.3) | 139(52.9) | 168(61.8) | 705(52.2) |
| The COVID-19 vaccine was developed too fast. | 240(87.3) | 141(53.0) | 132(49.4) | 234(82.1) | 160(60.4) | 907(66.8) |

Data shown as N (percent)

In terms of knowledge about the COVID-19 vaccine, 88.2% of the HCP indicated that the side-effects of COVID-19 vaccine are usually mild. About 66.8% of the respondents across all 5 countries reported that the vaccine was developed too fast. Additionally, 52% of the HCPs in all sites did not believe there was sufficient evidence that the COVID-19 vaccine prevents the occurrence and spread of COVID-19, with more than 60% of HCP in Burkina Faso and Ghana reporting this. About 17% of HCP believe that people had died after taking the COVID-19 vaccine (more than 22% in Ethiopia and Nigeria). Across all countries, a few respondents (< 10%) believed that it is not necessary to get COVID 19 vaccine if one follows all COVID-19 safety protocols (20% in Burkina Faso and 12% in Ethiopia). Less than 11% of all respondents believed the COVID-19 is a conspiracy or a bioweapon and 4% reported that people on the African continent are immune to COVID-19.

Almost all HCP from study countries perceived a high to very high risk of exposure to COVID-19 except in Burkina Faso, where about 41% of HCP perceived a low risk of exposure (Fig 2a). At least 28% of all HCP believed that the COVID-19 vaccines were very safe and up to 45% thought they were somewhat safe (Fig 2b). Only 1% of the HCP believed the COVID-19 vaccines were not safe at all. Approximately 8% believed they were not very safe, with the highest proportion with this belief in Ghana (21.0 %). The majority of respondents believed that COVID-19 vaccines were very or somewhat effective in preventing the disease. In Ethiopia 32% of respondents believed vaccines were not very effective (Fig 2c).

**Figure 2a:**
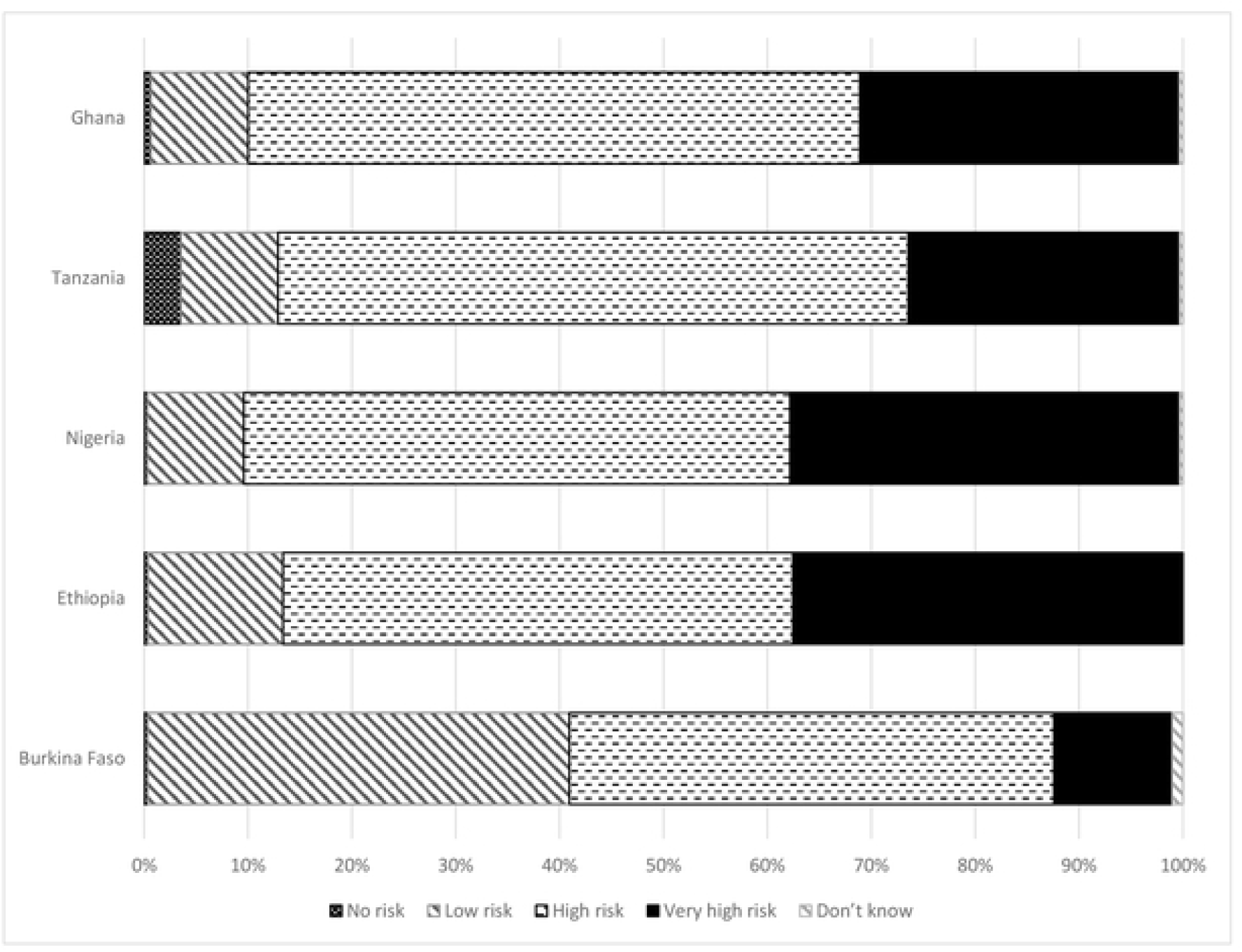
Health care provider perceptions of risk of COVID-19 exposure in 5 countries in sub-Saharan Africa.

**Figure 2b:**
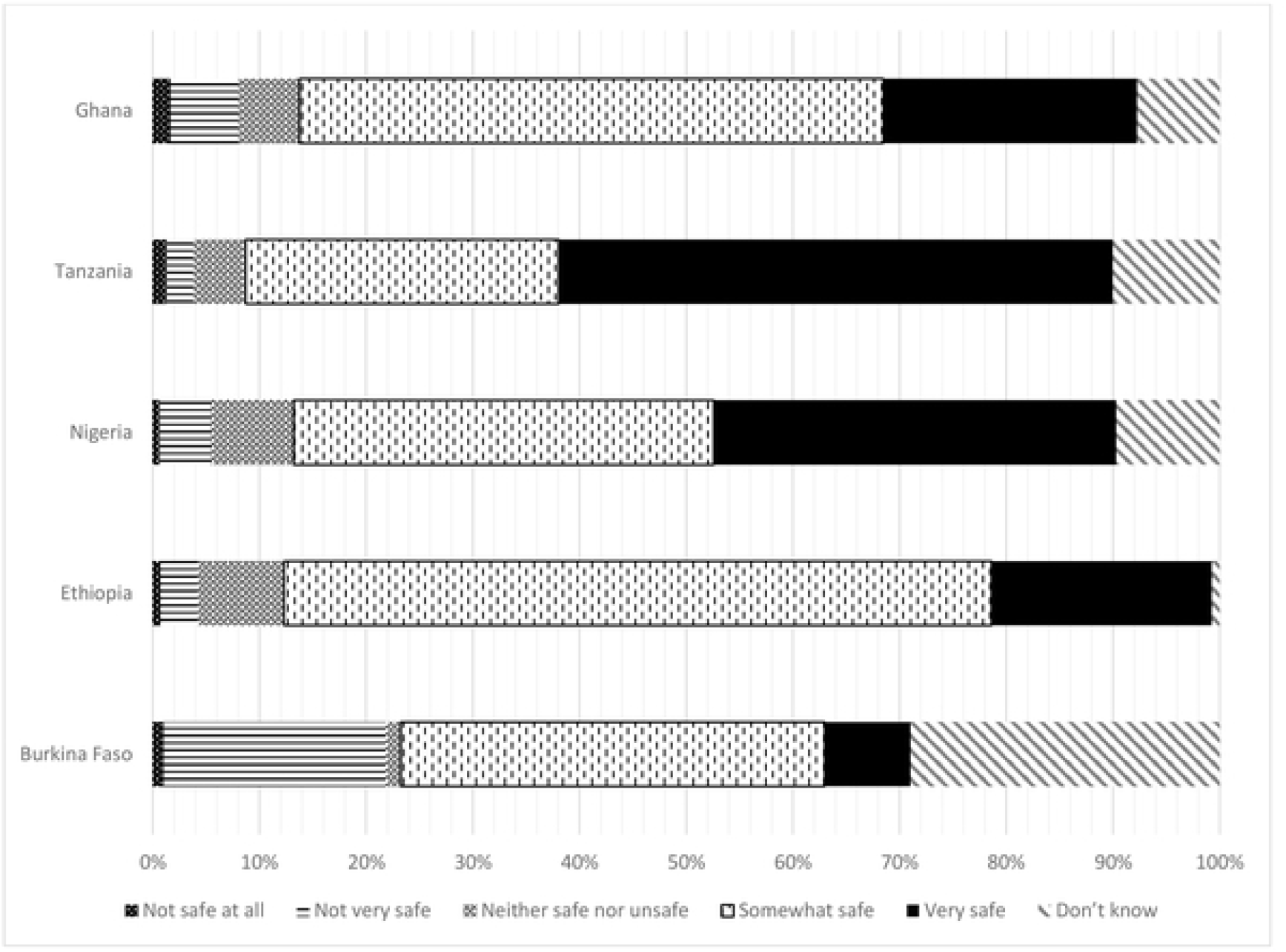
Health care provider perceptions on COVID-19 vaccine safety in 5 countries in sub-Saharan Africa.

**Figure 2c:**
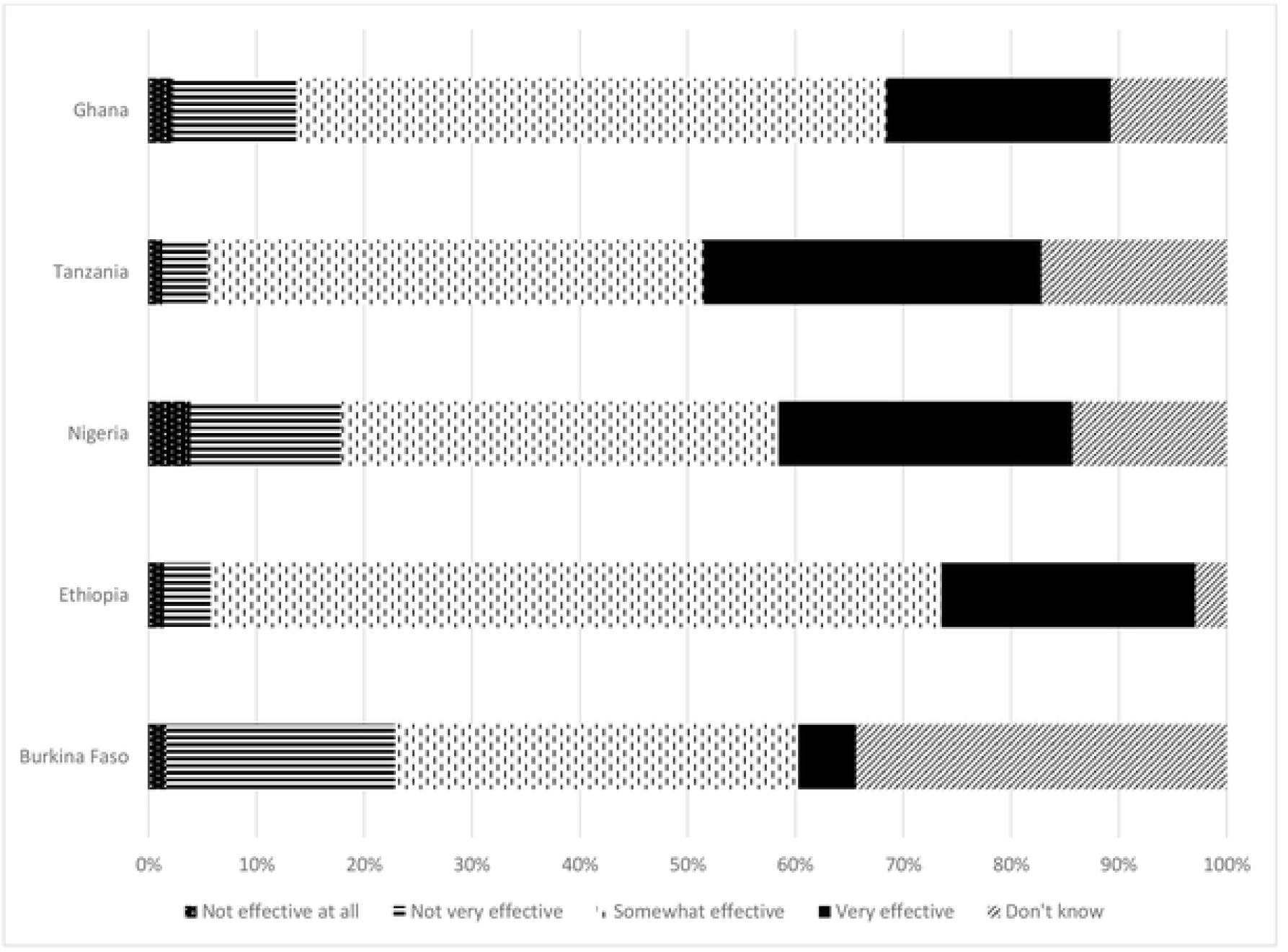
Health care provider perceptions on COVID-19 vaccine effectiveness in 5 countries in sub-Saharan Africa.

Table 3 shows beliefs about the COVID-19 vaccine, and willingness to take vaccines. At least 85% of the respondents indicated that COVID-19 vaccines were available in their country and localities, except in Ghana, where 45.7% reported vaccine availability at the time of the interview. The majority of HCPs (84%) indicated that COVID-19 vaccine was available to them as HCPs, with 67.9% of them having been vaccinated by the time of the survey, and a further 10.8% expecting to be vaccinated by the end of 2021. Vaccination rates were lowest in Burkina Faso (40.3%), Tanzania (66.5%) and Ghana (69.3%). At least 86% of HCP indicated that it was important for HCPs to get vaccinated. At least 77% of HCPs reported that their workplaces had formulated policies on COVID-19. Most participants from Nigeria (95.2%), Ghana (87.7%) and Ethiopia (75.1%) reported having workplace guidelines on COVID-19 but only 60.1% and 67.1% from Burkina Faso and Tanzania, respectively, had guidelines.

**Table 3:** Perceptions of the safety, effectiveness and availability of vaccines and willingness to get the COVID-19 vaccine and expected benefits in 5 countries in sub-Saharan Africa.

| Characteristics | Burkina Faso | Ethiopia | Nigeria | Tanzania | Ghana | Total |
| --- | --- | --- | --- | --- | --- | --- |
| **Perceived safety of vaccines** |  |  |  |  |  |  |
| In general, I believe that vaccines are safe | 153 (75.7) | 210 (76.6) | 292 (99.0) | 250 (89.3) | 279 (95.6) | 1183 (88.2) |
| **Vaccine hesitancy** | 137(45.7) | 27(9.8) | 25(8.1) | 79(25.7) | 27(9.0) | 295(19.7) |
| **Availability of COVID-19 vaccines** |  |  |  |  |  |  |
| Covid 19 vaccine available in country | 298 (100) | 237 (85.6) | 306 (98.1) | 306 (98.7) | 137 (45.7) | 1284 (85.8) |
| Covid 19 vaccine has been made available to you as a healthcare provider | 192 (64.4) | 258 (93.1) | 287 (92.0) | 299 (96.5) | 225 (75.0) | 1261 (84.2) |
| **Have you or any of your colleagues received the Covid 19 vaccination** |  |  |  |  |  |  |
| Yes, I have been vaccinated | 120 (40.0) | 226 (81.6) | 257 (82.4) | 206 (66.5) | 208 (69.3) | 1017 (67.9) |
| I know someone who has been vaccinated | 135 (45.0) | 29 (10.5) | 27 (8.7) | 97 (31.3) | 81 (27.0) | 369 (24.6) |
| **Workplace has formulated policies or guidelines related to COVID-19** | 179 (60.1) | 208 (75.1) | 297 (95.2) | 269 (87.7) | 208 (67.1) | 1161 (77.6) |
| **How important is it for Health care providers to get vaccinated against COVID-19** |  |  |  |  |  |  |
| Very important | 209 (70.4) | 217 (78.3) | 291 (93.3) | 284 (91.6) | 288 (96.0) | 1289 (86.2) |
| Somewhat important | 69 (23.2) | 49 (17.7) | 15 (4.8) | 19 (6.1) | 10 (3.3) | 162 (10.8) |
| Not very important | 11 (3.7) | 4 (1.4) | 2 (0.6) | 3 (1.0) | 0 | 20 (1.3) |
| Not important at all | 4 (1.4) | 6 (2.2) | 0 | 0 | 1 (0.3) | 11 (0.7) |
| Don't know | 4 (1.4) | 0 | 4 (1.3) | 4 (1.3) | 1 (0.3) | 13 (0.9) |
| **COVID-19 vaccine's country of origin affects my willingness to take the vaccine** | 74 (24.9) | 107 (38.6) | 56 (18.0) | 68 (21.9) | 91 (30.3) | 396 (26.5) |
| **If COVID-19 vaccine had been developed or tested in Africa would that affect your willingness to take it or recommend it** |  |  |  |  |  |  |
| No, will not change my mind | 88 (29.6) | 191 (69.0) | 198 (63.5) | 205 (66.1) | 209 (69.7) | 891 (59.6) |
| Yes, will decrease my chances of taking it | 2 (0.7) | 46 (16.6) | 41 (13.1) | 18 (5.8) | 26 (8.7) | 133 (8.9) |
| Yes, will increase my chances of taking it | 199 (67.0) | 32 (11.6) | 56 (18.0) | 68 (21.9) | 60 (20.0) | 415 (27.7) |
| **When do you think a COVID-19 vaccine will be made available to you** |  |  |  |  |  |  |
| Never | 4 (1.4) | 20 (7.2) | 3 (1.0) | 16 (5.2) | 2 (0.7) | 45 (3.0) |
| Already received the vaccine | 119 (40.1) | 226 (81.6) | 257 (82.4) | 206 (66.5) | 208 (69.3) | 1016 (67.9) |
| Before the end of 2021 | 88 (29.6) | 15 (5.4) | 21 (6.7) | 12 (3.9) | 26 (8.7) | 162 (10.8) |
| During the first six months of 2022 | 7 (2.4) | 9 (3.3) | 3 (1.0) | 7 (2.3) | 9 (3.0) | 35 (2.4) |
| During the last six months of 2022 | 2 (4.7) | 1 (0.4) | 3 (1.0) | 4 (1.3) | 2 (0.7) | 12 (0.8) |
| 2023 or later | 37 (12.5) | 4 (1.5) | 2 (0.6) | 9 (2.9) | 9 (3.0) | 61 (4.1) |
| Don't know | 39 (13.1) | 2 (0.7) | 21 (6.7) | 50 (16.1) | 44 (14.7) | 156 (10.4) |
| **Would you recommend that my friends and loved ones get the COVID-19 vaccine** | 57 (19.2) | 26 (9.4) | 11 (3.5) | 33 (10.7) | 6 (2.0) | 133 (8.9) |
| **Are you doing any activities to boost your community's confidence in taking the vaccine** |  |  |  |  |  |  |
| Community outreach/education | 214 (72.1) | 228 (82.3) | 185 (59.3) | 261 (84.2) | 193 (64.3) | 1081 (72.3) |
| Other | 0(0) | 0(0) | 23 (7.4) | 0(0) | 9 (3.0) | 32 (2.2) |

| Characteristics | Burkina<br>Faso | Ethiopia | Nigeria | Tanzania | Ghana | Total |
| --- | --- | --- | --- | --- | --- | --- |
| Willing to participate in a vaccine clinical trial if available | 181 (60.9) | 136 (49.1) | 102 (32.7) | 69 (22.3) | 141 (47.0) | 629 (42.1) |

Approximately 26.5% of HCPs indicated that the origin of the vaccine influenced their willingness to take COVID-19 vaccines, and 27.7% of all respondents indicated that if a vaccine were developed and tested in Africa, this would increase their willingness to take it. This was more so in Burkina Faso, where 67% of HCP indicated a greater willingness to take African vaccines. Among unvaccinated HCP, the majority indicated a willingness to take the vaccine if it was availed to them (Suppl Fig 1). However, in Tanzania, 48.1% of unvaccinated HCPs reported unwillingness and 18.3% were undecided on whether to take the COVID-19 vaccine. Reasons for unwillingness to take the COVID-19 vaccines across the countries include not thinking the vaccine was effective, concern about side effects, concern that vaccine development was too fast, fear of receiving poor quality vaccines and negative media reports. These concerns were most frequently reported in Burkina Faso (Fig 3). Finally, HCPs reported that the most prescribed treatments for COVID-19 were antibiotics/azithromycin (47.2%) and multivitamins (33.4%) (Supplementary Fig 2). Close to 20% of the cases were not prescribed any medications.

**Figure 3:**
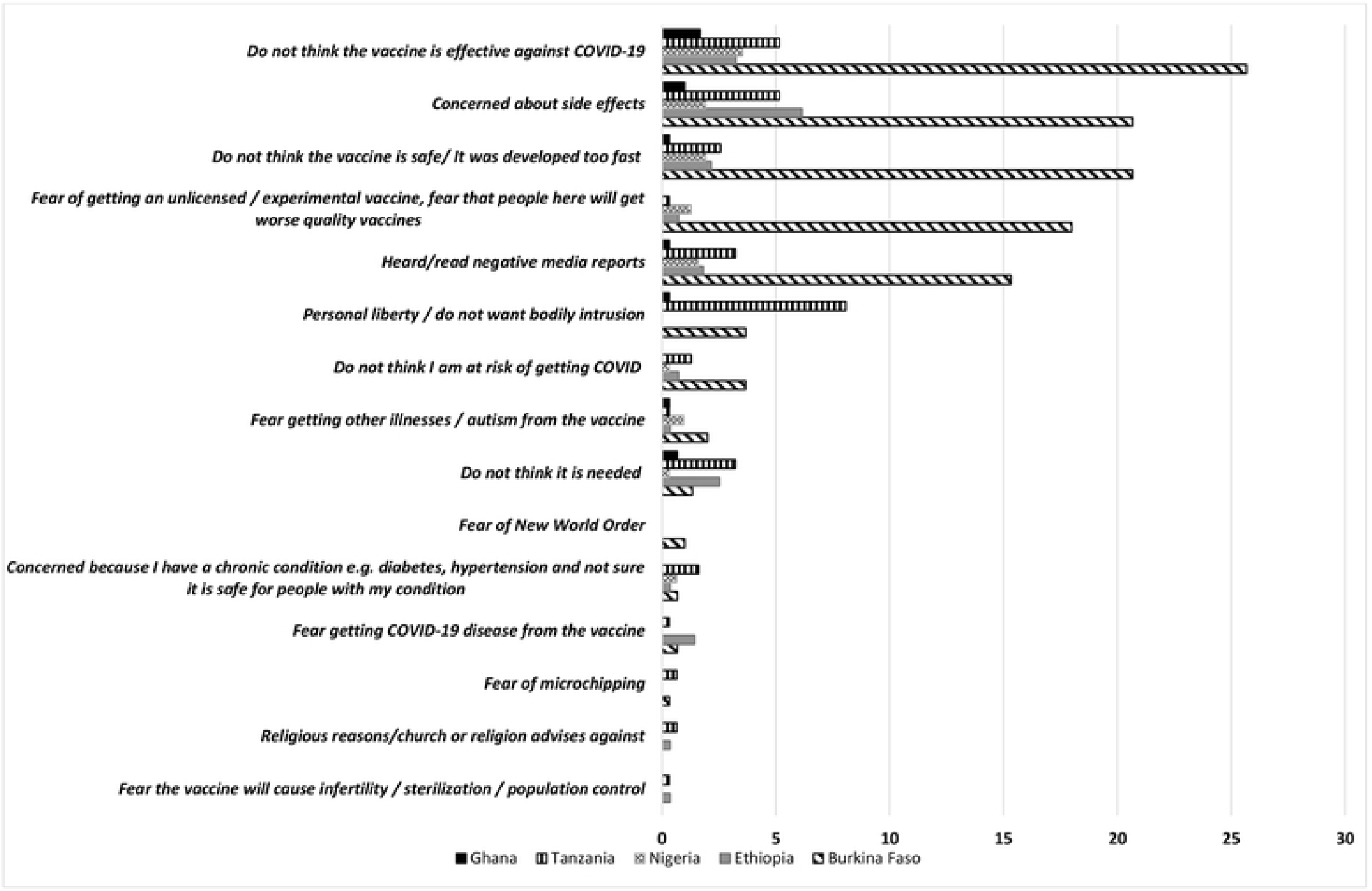
Frequent reasons (%) for not getting COVID-19 vaccination in 5 countries in sub-Saharan Africa.

COVID-19 vaccine hesitancy affected 19.7% of the HCPs (Table 3). Reported hesitancy was highest in Burkina Faso (45.7%) and Tanzania (25.7%) compared to other countries. The lowest levels of vaccine hesitancy were reported in Nigeria at 8.1% and Ghana at 9.0%.

Table 4 shows factors associated with COVID-19 vaccine hesitancy among HCP in the study. We found that there were site-specific differences in the risk of COVID-19 vaccine hesitancy among HCP. In Ethiopia (RR:0.52, 95% CI: 0.29,0.95) HCP had lower vaccine hesitancy, and in Tanzania, there was a trend towards having higher vaccine hesitancy (RR:1.49, 95% CI: 0.99,2.25, p=0.06) among HCP compared to Burkina Faso. Age was associated with vaccine hesitancy, with HCPs aged 45 years or older less likely to be vaccine-hesitant (RR:0.65, 95% CI: 0.44,0.95) compared to those aged 20-29 years. Nurses were more likely to be vaccine-hesitant (RR 1.38, 95% CI: 1.00,1.89) compared to doctors. Respondents reporting that COVID-19 vaccines are very effective (RR:0.21, 95% CI:0.08, 0.55) were less likely to report vaccine hesitancy. For each reason given by HCP for not wishing to receive COVID-19 vaccination, the risk of being vaccine-hesitant increased (RR 1.78, 95% CI:1.62, 1.95). Finally, HCP reporting that the World Health Organization or UNICEF endorsement of the COVID-19 vaccine as safe and effective would make them more likely (RR 0.51, 95% CI:0.35, 0.74) or much more likely (RR 0.69, 95% CI:0.52, 0.92) to receive the vaccine were less likely to be vaccine-hesitant.

**Table 4:** Factors associated with COVID-19 vaccine hesitancy among health care providers in round 2 of the ARISE COVID study in 5 countries.

|  | Unadjusted<br>RR (95% CI) | Adjusted<br>RR (95% CI) |
| --- | --- | --- |
| **Country** |  |  |
| Burkina Faso | Ref | ref |
| Ethiopia | 0.21(0.14,0.32) *** | 0.52(0.29,0.95)* |
| Nigeria | 0.18(0.12,0.27) *** | 0.67(0.37,1.23) |
| Tanzania | 0.56(0.43,0.74) *** | 1.49(0.99,2.25) |
| Ghana | 0.56(0.43,0.74) *** | 0.65(0.36,1.17) |
| Male sex | 0.86(0.68,1.09) | 0.90(0.69,1.18) |
| **Age** |  |  |
| <30 years | Ref | ref |
| 30-44 years | 1.19(0.92,1.54) | 0.80(0.59,1.08) |
| 45 years+ | 0.97(0.69,1.36) | 0.65(0.44,0.95)* |
| **Occupation** |  |  |
| Doctor | Ref | ref |
| Nurse | 1.18(0.90,1.54) | 1.38(1.00,1.88)* |
| Other occupation | 1.03(0.62,1.70) | 1.52(0.88,2.63) |
| **Religion** |  |  |
| Catholic or none | Ref | ref |
| Muslim | 3.10(2.10,4.60)*** | 1.00(0.58,1.72) |
| Orthodox Christian | 3.18(2.13,4.74) *** | 1.16(0.68,1.99) |
| Protestant or other | 1.98(1.33,2.95) ** | 1.16(0.71,1.89) |
| **Health centre type** |  |  |
| Government hospital/clinic | Ref | ref |
| Private hospital/clinic | 1.12(0.42,1.51) * | 0.96(0.70,1.33) |
| Health outpost/CHPS compounds or other | 0.58(0.41,0.84) ** | 0.73(0.47,1.13) |
| Perceived risk of COVID-19 disease |  |  |
| Low risk | 1.30(0.48,3.53) | 0.94(0.33,2.69) |
| No risk | 0.65(0.24,1.75) | 0.93(0.33,2.61) |
| Very high risk | 0.58(0.21,1.59) | 1.08(0.38,3.07) |
| High risk | Ref | ref |
| Effectiveness of COVID-19 vaccine |  |  |
| Very effective | 0.05(0.02,0.12)*** | 0.21(0.08,0.55)* |
| Somewhat effective | 0.31(0.18,0.53)*** | 0.87(0.46,1.63) |
| Not very effective | 0.91(0.52,1.58) | 0.97(0.53,1.80) |
| Not effective at all | Ref | ref |
| Workplace policy score (number of policies) | 0.75(0.68,0.82)*** | 0.98(0.88,1.09) |
| Reasons for not vaccinating (number of reasons) | 2.03(1.90,2.16)*** | 1.78(1.62,1.95)*** |
| Believe the COVID-19 vaccine is a bioweapon | 2.89(1.99,4.20)*** | 0.94(0.62,1.42) |
| Believe vaccines are safe | 0.30(0.22,0.40)*** | 0.71(0.50,1.02) |
| WHO/UNICEF endorsement of the COVID-19 vaccine as safe and effective affects likelihood get the vaccine |  |  |
| Much more likely | 0.19(0.13,0.26)*** | 0.51(0.35,0.74)*** |
| More likely | 0.40(0.31,0.52)*** | 0.69(0.52,0.92)* |
| No difference | Ref | ref |
| Less likely | 0.50(0.27,0.90)* | 0.92(0.47,1.83) |
| Much less likely | 0.84(0.31,2.29) | 1.53(0.51,4.56) |

376 Acronyms: COVID-19; Coronavirus disease 2019, RR: relative risk, CI: Confidence interval, \* <0.05, \*\*<0.01, \*\*\*<0.001

## DISCUSSION

We assessed perceptions and predictors of COVID-19 vaccine hesitancy among HCP in Burkina Faso, Ethiopia, Ghana, Nigeria and Tanzania. We found that the majority of HCP had been vaccinated by the time of the survey and that almost a fifth of the HCPs across the sites were hesitant to receive the COVID-19 vaccine. There were differences in vaccine hesitancy by site. The age of a HCP, their profession and perceptions about COVID-19 vaccine effectiveness were significant predictors of vaccine hesitancy in this study. Older HCP reported lower levels of vaccine hesitancy, while nurses and HCP who believed that COVID-19 vaccines were not effective were more likely to be vaccine-hesitant.

Our findings show consistency with some findings from previous studies. In a study of vaccine hesitancy among HCPs in South Africa, being an older HCP or a physician and perceptions of the benefits and risks of vaccination were associated with lower vaccine hesitancy [13]. However, the study also found additional factors that influenced vaccine hesitancy among HCPs including beliefs that vaccines are incompatible with religion and willingness to be vaccinated to protect others [13]. Similarly, another study in Ethiopia found that older age (>40 years) and being a medical doctor were associated with a lower risk of vaccine-hesitancy. In Nigeria, a study found that HCP who were younger, single and with lower income had more vaccine hesitancy, while being a nurse or doctor was associated with lower hesitancy [17]. In Ghana, being female and having a low income have also been associated with increased COVID-19 vaccine hesitancy [18]. These findings suggest there may be information asymmetry for example among different cadres of HCPs and lower perceived risks among some population sub-groups (20). Women additionally face greater concerns about the effects of COVID-19 vaccines on their fertility (21). In this study, we did not observe greater hesitancy among female HCP.

Older age may be associated with lower vaccine hesitancy since age is a risk factor for COVID-19 infection, and that older people are more vulnerable to more severe disease possibly due to more co-morbidities. This could be a factor influencing decisions by older HCPs to be vaccinated. In an Africa-wide survey, respondents indicated that vaccine hesitancy was lower among those with a higher perceived risk of infection [19]. Additionally, HCP who perceive that vaccines are effective may face fewer personal barriers and may be more willing to take the vaccine. Other factors reported by studies include concerns about vaccine safety, serious side effects and efficacy of vaccines, with limited information also being a key factor (16). Additionally, perceptions of lack of COVID-19 vaccine benefits, distrust of the government and the ability of science to provide safe and effective vaccines and concerns about vaccine safety were associated with greater vaccine hesitancy [14].

The differences between our findings and those from previous studies may be because we included 5 countries in our study and have a larger sample size and variability of responses than previous studies. Our findings are able to account for differences in contextual factors across the countries and determine overall influential factors affecting vaccine hesitancy across countries.

In our study, there were differences in vaccine hesitancy among HCP by country, with Burkina Faso and Tanzania reporting the highest levels of hesitancy and Ghana and Nigeria showing low levels of hesitancy. Previous studies have also shown variations in acceptance rates from as low as 28% in Central Africa, and up to 48-49% in West and East Africa (16), suggesting that there may be context-specific factors influencing attitudes towards COVID-19 vaccines. Country-specific factors such as differences in availability and access to vaccines, the burden of COVID-19 cases, and variation in mitigation measures put in place could account for differences across countries. Reported rates of vaccine hesitancy have been higher in other studies. A study conducted early in the COVID-19 emergency in Nigeria found higher rates of vaccine hesitancy among HCPs (50%) [17]. In a South African study, 41.0% of HCPs were vaccine-hesitant [13] and in Ghana, three out of five HCP were hesitant early in the pandemic (22). Levels of vaccine hesitancy of 41%-45.9% have also been reported among HCP in South Africa and Ethiopia [7]. These studies were conducted early during the COVID-19 pandemic before vaccines had been available. Additionally, low rates of COVID-19 vaccine acceptance by the general population on the African continent have been reported [20]. Our observations of lower levels of vaccine hesitancy may be partly influenced by changing attitudes and beliefs towards the COVID-19 vaccine across the continent as the pandemic has progressed, due to increased cases and mortality, increased information and greater availability of COVID-19 vaccines.

We observed higher levels of COVID-19 vaccination among HCP across our study, compared to previous reports on the African continent. Two-thirds of HCP interviewed across the 5 countries had already received the COVID-19 vaccination at the time of the survey; however, vaccination rates were low in Burkina Faso. Health care providers were prioritized to receive the COVID-19 vaccine initially when the vaccines were not readily available. It is therefore expected that more HCP had received the vaccine. The WHO, however, reported that approximately 25% of HCPs were fully vaccinated against COVID-19 on the African continent by November 2021 [8]. The differences observed between this study and previous studies could be due to the fact our study is more recent, and vaccines may have been relatively more available during our study. Additionally, we only assessed vaccination status in a few selected health centers in the study sites, and our estimates are not nationally representative. Further, four of our survey sites were in urban areas, where vaccination rates are higher. Reasons for lower vaccination in some of our study sites could have been due to challenges with the availability of the COVID-19 vaccines, and this is consistent with the situation across the African continent. Reasons for the limited availability of COVID-19 vaccines on the African continent have included global architecture and restricted vaccine supply chains and limited availability of donations promised by donors through the COVAX facility for low- and middle-income countries by the end of 2021 [21].

Another possible reason for the higher vaccination rates and lower vaccine hesitancy we observed in our study could be because perceptions about the COVID-19 vaccine were mainly positive in our study. There was a high willingness among the unvaccinated to get vaccinated across three of our five sites. While some COVID-19 misinformation was reported, the proportion of participants reporting misinformation was low, with few HCPs believing that COVID-19 is a conspiracy or bioweapon; that people of African descent are immune to COVID-19; that people have died from taking the vaccine; and that the COVID-19 vaccine was unnecessary. Many in the study correctly believed that the side effects of the COVID-19 vaccine are usually mild and temporary. The low levels of misinformation overall could have contributed to lower levels of hesitancy among HCPs.

While most of the HCP who had not been vaccinated were willing to take the COVID-19 vaccine, in Tanzania, willingness to take the COVID-19 vaccination was low. The main reasons for this were perceptions that the vaccine was not effective, concerns about side effects and the belief that the vaccine was developed too fast. The concerns about side effects and safety have also been reported in other studies from similar settings and among the general population (20, 23, 24).

Beliefs regarding the safety and effectiveness of the COVID-19 vaccine are important considerations for HCP uptake. The majority of HCP in this study believed that COVID-19 vaccines are safe and effective in preventing the disease. In this study, HCP rightly perceived a high risk of exposure to COVID-19 across all countries, an observation likely due to HCP attending to patients which puts them at a higher risk of exposure to the disease. In contrast, most of the HCP believed that there is insufficient evidence that the COVID-19 vaccine prevents the occurrence and spread of COVID-19. This observation could reflect sentiments that although Europe, America and Asia had greater availability of vaccines, their reported declines in morbidity lagged as vaccine rollout continued in these contexts. Finally, there were concerns about how quickly the vaccine had been developed in this study since drug development usually takes many years to go through the various pre-clinical and clinical stages. In addition, at least a quarter of respondents indicated that the origin of the vaccine influenced their willingness to take COVID-19 vaccines, and a similar proportion indicated that if a vaccine were developed and tested in Africa, this would increase their willingness to take it. Potential concerns about vaccine safety and efficacy may be partially addressed in the future by encouraging local production of vaccines on the African continent.

Our study had limitations. Being a telephone survey, it would not have been equally available to all health workers, and it was affected by non-response in some countries. We instituted replacements using randomly allocated individuals from existing sampling frames and HCP lists to mitigate against this. Additionally, 4 of our 5 study sites were urban, and therefore findings may not be attributable to HCPs in rural areas.

In conclusion, in this study, we found low COVID-19 vaccine hesitancy across most countries, with higher vaccine hesitancy among HCP in Burkina Faso and Tanzania. We also found that factors associated with hesitancy include age, occupation, beliefs about vaccine effectiveness and endorsement by global public health institutions. Efforts to increase vaccine uptake will have to address vaccine hesitancy by tackling information and knowledge gaps among different cadres of health workers, along with efforts to increase vaccine supply.

## Data Availability

Individual participant data collected for this study may be shared based on a reasonable request to the corresponding authors.

## Abbreviations

ARISE African Research: Implementation Science and Education network
COVID-19: Coronavirus disease 2019
HCP: Health Care Providers
PPE: personal protective equipment
SSA: sub-Saharan Africa

## Acknowledgments

We thank all study participants and data collectors for contributing to this study. The survey team in Ghana appreciates support from the Kintampo Health Research Centre of Ghana Health Service, and community leadership of Kintampo North Municipality and Kintampo South District. We acknowledge institutional support from Harvard T.H. Chan School of Public Health, Boston, MA; Harvard University Center for African Studies, Boston, MA; Heidelberg Institute of Global Health, Germany and the George Washington University Milken Institute of Public Health, Washington, DC who provided support for the work.

## Authors’ contributions

IM and LNA conceived and designed the study, analyzed the data, and drafted the manuscript. WWF was the principal investigator for the parent study, conceived the study, designed the study, interpreted the data, and guided revisions of the manuscript. EP, TA, VB, CJ, DW, FM, OM, NA,AC, FW, BL, ECH, AI, SA, KPA,YB, JK, AO, AS, AS, SV, MMS, SV, ES, TB and RT designed the study, interpreted the data and revised the manuscript. All authors read and approved the final manuscript.

